# Mortality of individuals with antemortem genetic testing for *PRNP* variants in the United States, 1998-2024

**DOI:** 10.1101/2025.10.03.25337271

**Authors:** Yuan Lian, Keisi Kotobelli, Kathleen Glisic, Daniel A Sprague, Sonia M. Vallabh, Brian S. Appleby, Eric Vallabh Minikel

## Abstract

**Objectives:** To characterize the survival of individuals with pathogenic *PRNP* variants — including to estimate annual hazards, to judge the accuracy of previously reported survival data, and to evaluate the utility of public record searches in determining vital status.

**Methods:** In this single center cohort study, we gathered data on individuals who received positive antemortem *PRNP* genetic tests at the U.S. National Prion Disease Pathology Surveillance Center (NPDPSC). Genetic test results and autopsy status were queried from the NPDPSC database, and public record searches were conducted using online tools.

**Results:** 404 individuals received positive genetic test results. Of 206 cases symptomatic at the time of genetic testing, 188 are likely now deceased based on typical disease duration for their genetic variants. Combined autopsy and public record searches in combination confirmed 174 of these deaths, for an estimated sensitivity of 92.6%. Of 111 autopsied individuals, evidence of death was found in public record searches for 109, a sensitivity of 98.2%. Of 198 individuals who were asymptomatic at the time of testing, 32 since died of definite or possible prion disease. Among 20 of these who underwent autopsy, public record searches confirmed deaths of 18, for a sensitivity of 90%. Among 99 E200K individuals over 936 person-years of follow-up, 18 deaths were observed, significantly fewer than 27.4 expected according to life tables based on retrospective data. The age-dependent penetrance of E200K calculated from these longitudinal data was significantly lower than that from retrospective data, with 69% penetrance by age 80 and a median age at death of 75. No significant difference was found for D178N, which appears highly penetrant, though the median age at death was numerically higher than seen in retrospective data. V210I was associated with just 2 deaths, both after age 90, consistent with minimal penetrance.

**Discussion:** These data support the accuracy of penetrance classifications for *PRNP* variants reported based on retrospective data, but may suggest an age of onset distribution shifted slightly later than that calculated retrospectively. Autopsy data and public death records in combination were sensitive and concordant, but additional prospective data should be gathered to support future preventive trials.

## Introduction

Prion disease can be caused by genetic variants in *PRNP*, only some of which are highly penetrant^1^. Most data on age at symptom onset and penetrance have been gathered chiefly from reports of deaths to surveillance centers^2^. Hazards estimated from these data could be inaccurate if penetrance is incomplete or if reporting is biased. Accurate estimation of annual risk of disease onset or mortality for at-risk individuals may be important for clinical development of drugs intended to delay disease onset. Here, we examine a cohort of individuals who received a positive antemortem *PRNP* genetic test. We evaluate the utility of public record searches to determine their current vital status, and we estimate annual hazards for individuals who were asymptomatic at the time of genetic testing.

## Methods

### Standard protocol approvals, registrations, and patient consents

This study was approved by University Hospitals Cleveland Medical Center’s Institutional Review Board (IRB) covering de-identified data of discarded specimens used in surveillance activities (01-14-18). IRB provided a waiver of informed consent. This study was also approved by the Broad Institute’s Office of Research Subjects Protection (NHSR-2121).

### Study participants

Participants were N=404 individuals with blood DNA testing (221 female, 183 male) performed at the National Prion Disease Pathology Surveillance Center (NPDPSC) from December 1998 through January 2024 for whom a *PRNP* variant was identified.

### Public record searches

The patient’s full name, date of birth, and/or state of residence were searched using Google during April 2024 - August 2025 to verify their mortality status through obituary listings as well as other websites including death announcements and cemetery records. For individuals for whom records were not found through these sources, the Social Security Death Index (SSDI) was searched for additional verification of mortality status.

### Life tables

We filtered previously reported life tables^2^ to include only individuals who had not been followed prospectively, but rather, whose vital status was only known as a single moment snapshot due to presentation to a clinical center, report to a surveillance center, or ascertainment in a family history collected from a related proband. These data were not considered to be left-truncated or left-censored (all individuals were assumed to have observation time beginning at age 0), but were considered right-censored (where applicable) at the time an individual was last known to be alive. For antemortem genetic testing data, we considered the NPDPSC data to be both left-censored at the time that individuals underwent predictive testing, and right-censored (where applicable) at the time they were last known to be alive. These assumptions and methodology have been previously described in detail^2^. Briefly, for each time interval (integer ages from 1 to 100), we calculated the number of lives under observation (Lt), deaths (Dt), and censorings (“withdrawals”, Wt), Lives under observation (Lt) was calculated as the number of individuals who underwent genetic testing at or before age t, *and* whose death or censoring occurred at or after age t. The fact that each individual increments Lt only at the age when they undergo genetic testing accounts for delayed entry. Deaths (Dt) was calculated as the number of individuals whose death occurred at age t. Censorings (Wt) was calculated as the number of individuals for which no evidence of death was found, therefore presumed to be still alive, and currently age t. The hazard, or probability of onset, at each age (Qt) was then calculated as Dt/Lt, and the proportion surviving (Pt) was calculated as the cumulative product of 1-Qt across all observed ages up to age t. For any ages where Lt was 0, (in particular, note that ages <18 are not included in this study) we assumed Qt to be 0. Median survival was calculated as the first integer age at which Pt drops below 50%. Penetrance at age 80 was calculated as the Pt at age 80.

### Statistical analysis

Sensitivity statistics utilized Wilson binomial confidence intervals. 90th percentiles of disease duration (1.4 years for E200K and 2.1 years for D178N, see Supplement for additional variants) were calculated from previously published disease duration tables^2^, using the number of months after disease onset when survival drops to 10%. Probabilities of onset per individual were calculated by using the smoothed annual hazards from the snapshot life tables described above. The Cox proportional hazards model for E200K was fit using left and right truncation times, with source dataset as the independent variable. All survival statistics (medians and proportions surviving at specific ages) and all data visualizations of NPDPSC data are computed accounting for left truncation as described under “Life tables” above. All analyses were conducted using custom scripts in R 4.2.0.

### Data availability

The full de-identified dataset will be made available to qualified investigators with ethical approval and a data use agreement upon request. Binned life tables are provided in the Supplementary Materials. These life tables and summary statistics are freely available at github.com/ericminikel/npdpsc_prospective

## Results

### Demographics and mortality

We identified N=404 individuals with positive antemortem genetic tests for whom the disease status at the time of genetic test was known (Figure S1). Age at test was (mean±SD) 52.8±11.9 for symptomatic (N=206) and 42.9±16.1 for asymptomatic (N=198) (Table 1). Of these individuals, N=131 went on to autopsy at the NPDPSC; for an additional N=4, NPDPSC was notified of the death but autopsy was not performed. Searches identified public death records for an overlapping set of N=205 individuals (online obituaries, N=173; Social Security death index, N=20; other websites N=12). Most (N=126, 61.5%) such records provided no information as to cause of death, with just N=16 (8%) providing specifics identifying prion disease as the cause of death, N=19 (9%) alluding to an illness without specifics, and N=44 (21.5%) calling for memorial donations to a prion disease-related organization.

**Table 1.** Concordance of autopsy and public record search results. Contingency table of individuals with blood DNA testing by disease status at time of test, autopsy status at NPDPSC, and results of public record searches.

| Status at time of genetic test | Death known from autopsy | Death discoverable from public records |  | Totals |
| --- | --- | --- | --- | --- |
|  |  | No | Yes |  |
| Symptomatic | No | 32 | 63 | 95 |
|  | Yes | 2 | 109 | 111 |
| Asymptomatic | No | 163 | 15 | 178 |
|  | Yes | 2 | 18 | 20 |
| Totals |  | 199 | 205 | 404 |

### Sensitivity of autopsy data and public record searches in symptomatic individuals

Among patients who were symptomatic at the time of genetic testing and had gone to autopsy, public records documented the deaths of 109/111, for a sensitivity of 98.2% (95%CI: 93.7-99.5%). Public records also indicated the deaths of 63 additional symptomatic individuals who had not been autopsied. We further examined the data for the remaining 32 symptomatic patients for whom neither autopsy nor public records indicated death. Of these 32 individuals, 6 had rare genetic variants for which disease duration is not well characterized. For another 12, the time elapsed since genetic testing was less than the 90th percentile of disease duration for their respective genetic variants, making it plausible that these individuals are still alive. The remaining 14 individuals were past the 90th percentile of disease duration for their respective mutations, making it very likely they are in fact deceased. If so, then a total of 188 individuals symptomatic at the time of genetic testing are deceased, and autopsy plus public record searches in combination detected 174 of these deaths, for a sensitivity of 92.6% (95%CI: 87.9-95.5%).

### Disease onset and mortality in initially asymptomatic individuals

Among individuals asymptomatic at the time of genetic testing, public records were found for 18/20 autopsied individuals (Table 1), for a sensitivity of 90% (95%CI: 69.9-97.2%). Autopsy confirmed prion disease as the cause of death in 17/20 individuals, while in 3 instances (all E200K), autopsy of an individual who had never developed prion disease symptoms confirmed that prion disease was not the cause of death. Public records also identified an additional 15 deaths, but generally did not state whether the individual had died of prion disease. In total, of 198 asymptomatic persons during 1,836 person-years of follow-up, 35 deaths occurred, with 32 deaths being either definitely (N=17) or possibly (N=15) related to prion disease (Table 2).

**Table 2.** Deaths and person-years of follow-up by genetic variant among individuals asymptomatic at time of testing. High vs. low penetrance classifications are based on literature evidence for Mendelian segregation or de novo status reported previously^1^. Deaths observed excludes the 3 autopsied individuals known to have died without developing prion disease. Mean±SD values are raw values computed from the data; because these statistics do not refer to survival, they are not calculated from a model accounting for left truncation.

| Estimated penetrance | Genetic variant | N individuals | Follow-up (person-years) | Mean±SD age at genetic test | Mean±SD years of follow-up | Deaths observed |
| --- | --- | --- | --- | --- | --- | --- |
| high | P102L | 5 | 43 | 28.2±8.7 | 8.6±3.0 | 0 |
|  | D178N | 31 | 295 | 37.2±12.8 | 9.5±4.4 | 6 |
|  | E200K | 99 | 936 | 46.9±15.9 | 9.5±5.6 | 18 |
|  | Other | 25 | 269 | 33.6±14.3 | 10.8±5.1 | 4 |
| low | V210I | 26 | 187 | 45.8±18.0 | 7.2±4.0 | 2 |
|  | Other | 12 | 106 | 44.0±12.2 | 8.8±5.2 | 2 |
| **Total** | - | 198 | 1,836 |  |  | 32 |

### Estimation of annual hazard in asymptomatic individuals

The probability of disease onset in each year of life, or annual hazard, has previously been estimated from “snapshot” data, primarily based on presentations of newly symptomatic cases to surveillance and clinical centers. We sought to estimate the annual hazard from longitudinal follow-up of initially asymptomatic individuals in our genetic testing cohort, and compare these longitudinal data to the previously reported snapshot data. We focused on D178N, E200K, and V210I, both because the number of person-years of longitudinal follow-up was largest, and because these variants are rapidly progressive, with disease duration typically <1 year, thus, age at onset and age at death are nearly identical^2^. For more slowly progressive variants, the available snapshot data capture age at symptom onset^2^, which is a poorer proxy for the age at death captured here by autopsy and public record searches.

From previously reported snapshot data, we extracted life tables for E200K and D178N individuals who had not been prospectively followed, and derived survival curves (solid lines, Figure 1A-B). We then produced life tables from our longitudinal data for asymptomatic individuals (dashed lines, Figure 1A-B), making the following assumptions: individuals whose deaths were not identified in autopsy data nor in public record searches were presumed to be alive at their current age; individuals where autopsy ruled out prion disease were censored at their age of death; individuals with public records only were presumed to have died of prion disease. As a comparator, we also plotted the lifespan of the general population based on the 2022 Social Security actuarial life tables (gray curves, Figure 1A-C).

**Figure 1.**
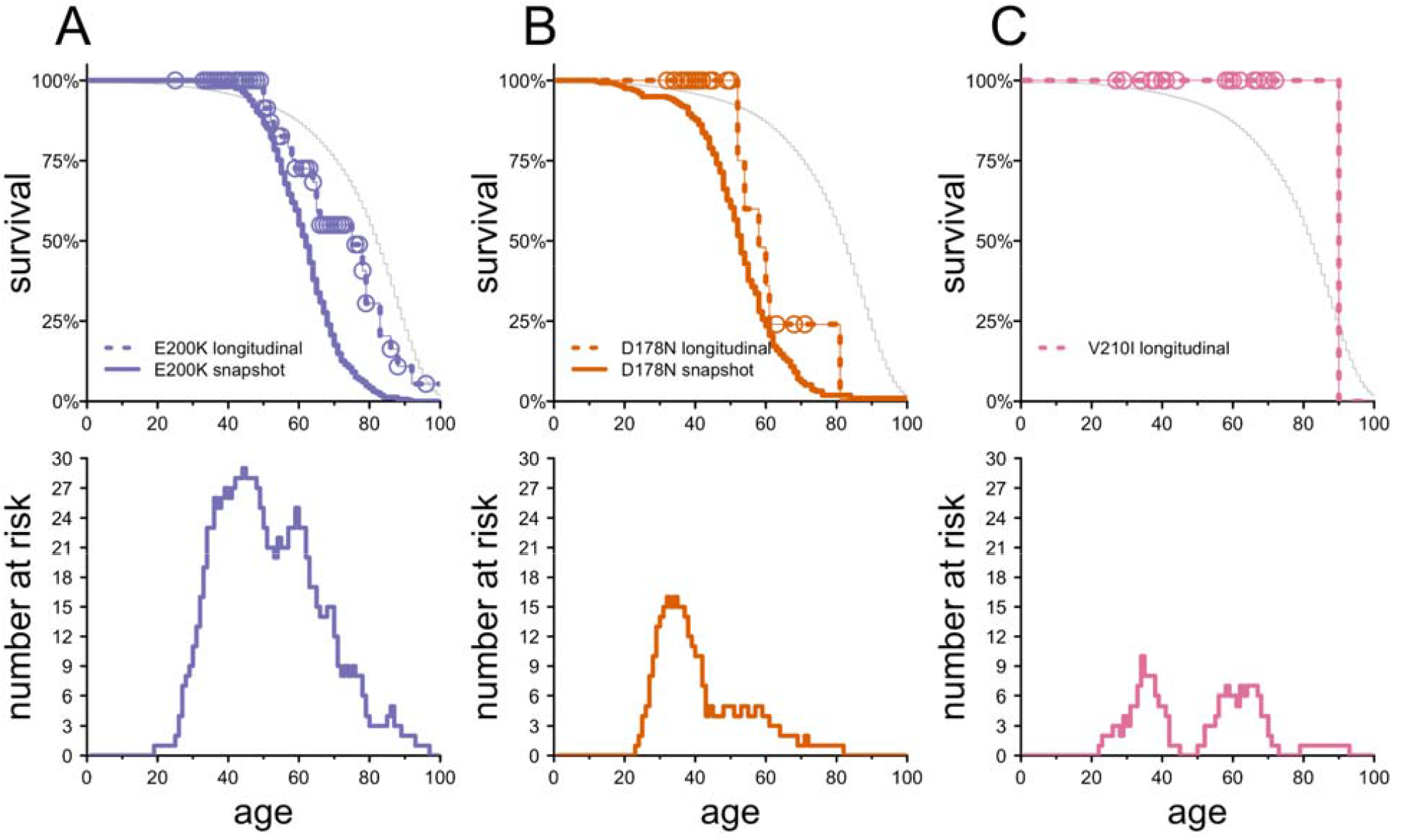
Survival of E200K, D178N, and V210I individuals by age. **A)** E200K, **B)** D178N, and **C)** V210I survival as a function of age. The top panels show survival curves (dashed lines) based on longitudinal NPDPSC asymptomatic genetic testing data, with censored datapoints (individuals alive at a particular age) shown as open circles. For E200K and D178N, the snapshot survival curve is superimposed as a thick solid line for comparison; for V210I, no snapshot data are available. Bottom panels show the number of individuals at risk in the NPDPSC asymptomatic genetic testing data.

For E200K, the longitudinal data survival curve of initially asymptomatic individuals was significantly right-shifted compared to the snapshot survival curve (P = 0.00011, Cox proportional hazards model). The longitudinal and snapshot datasets respectively yielded median ages at death of 75 versus 62, and penetrance by age 80 of 69% versus 96%. We tested the sensitivity of these estimates to our assumptions that deaths discovered through public record searches were due to prion disease, and that individuals without any evidence of death are alive today. We tested the most extreme alternative assumptions: i) that none of the 4 deaths of E200K individuals discovered via public record searches were due to prion disease, and ii) that 3 deaths due to prion disease were missed (a missing rate corresponding to the lower bound of the confidence interval of our sensitivity). These alternative assumptions would provide median ages of onset of 66 to 78 and penetrance by age 80 of 66% to 75% (Figure S2). In either extreme, the data would still classify E200K as highly penetrant but with a later-shifted age of onset distribution than previously reported. For D178N, any discrepancy between the longitudinal and snapshot data appeared smaller (median survival 58 vs. 52) and was non-significant (P = 0.21, Cox proportional hazards model). For V210I, no snapshot data were available, but in the longitudinal data (Figure 1C), only 2 deaths were observed, both over age 90 and with no indication of prion disease based on the public records found, consistent with low penetrance.

We also examined other possible confounders and explanations for the differences between our longitudinal and snapshot results. Even when the longitudinal dataset is analyzed using naive methods, without accounting for censoring or for delayed entry into the dataset at the time of genetic testing, median ages of onset for D178N and E200K remain higher, and penetrance at age 80 lower, than in the snapshot dataset (Table S1). This phenomenon is expected given that individuals only enter the longitudinal dataset at their age of genetic testing, and are thus depleted for younger disease onsets, an ascertainment bias that is correctly addressed by the analysis approach used throughout this paper. We also examined whether under-ascertainment of prion disease in older individuals in the snapshot dataset could explain the differences observed here (Figure S3). Even under the most extreme assumption of needing to correct for 75% under-ascertainment at age 60+, the median disease onset for E200K in the snapshot data would rise only from 62 to 65, smaller than the difference observed in the longitudinal data. Finally, we asked whether genetic testing prompted by initial symptoms could bias survival downward in the longitudinal dataset. Of the 24 deaths for D178N and E200K observed in our longitudinal dataset, only 3 occurred within the 90th percentile of disease duration for their respective genetic variants, while the remaining 21 observed deaths occurred after enough time had passed that it is unlikely the genetic testing was prompted by any suspicion of symptoms (see Discussion).

## Discussion

The penetrance and age of onset of *PRNP* variants have previously been classified based on the presence of Mendelian segregation and/or de novo variation in the literature, comparison of allele frequency in cases and controls, and aggregation of age of onset data chiefly as “snapshots” of symptomatic or deceased cases reported to surveillance centers worldwide. All of these methods are useful but are imperfect and subject to ascertainment bias. Individuals are generally more likely to appear in datasets when they develop disease (as opposed to never becoming sick), and are diagnosed, which may be a function of their age (better ascertainment in younger individuals) and their year of onset (improvements in surveillance and diagnosis over time^3,4^). Ascertainment bias can sometimes lead to surprising results, such as the false appearance of genetic anticipation^5,6^.

The ascertainment bias in snapshot data, wherein individuals are usually discovered only upon presentation with active disease, is difficult to correct for, because the number of individuals who either never presented symptoms, or were not diagnosed, cannot be empirically measured. A more ideal way to determine the age-dependent penetrance of genetic variants would be to longitudinally follow individuals with disease-causing variants and determine the proportion that succumb to disease in each year of life. This method carries its own ascertainment bias, specifically, that individuals only enter the dataset at the age at which they undergo genetic testing, but importantly, our method corrects for this bias in delayed entry to the at-risk population.

Herein we examined the survival of individuals with pathogenic *PRNP* variants identified when still asymptomatic, capturing 1,836 person-years of follow-up on 198 individuals.

The *PRNP* E200K variant is generally believed to cause genetic prion disease with high penetrance, based on its segregation with disease in many multigenerational families, the high proportion of cases with a positive family history, and its dramatic enrichment in prion disease cases over controls^1,7,8^. Most reports on E200K have estimated >90% penetrance by age 80^2,5,9,10^. 2 reports argued for a lower penetrance, ∼60%, but in small cohorts lacking any public dataset or any detailed description of the methods used for ascertainment or analysis^11,12^. Among 99 E200K individuals examined in the longitudinal dataset here, 18 died of definite or possible prion disease, with a median survival of 75 and estimated 69% penetrance by age 80. This differed significantly from the snapshot survival data, which would have predicted 27 deaths among the same individuals, with median survival of 62, and 96% penetrance by age 80. Our data still support classification of E200K as highly penetrant, but may point to a slightly more optimistic distribution of ages of onset.

The D178N variant has an earlier age of onset and an even higher proportion of cases with positive family history, perhaps suggesting less opportunity for ascertainment bias to cloud the snapshot data. Indeed, there was no significant difference in the survival of D178N individuals in our dataset compared to snapshot data: the median survival was slightly later — 58 versus 52 — but given the small number of deaths (6 deaths observed compared to 7 expected), this was within sampling variance. Our data support classification of D178N as highly penetrant.

Families with V210I generally do not exhibit multiplex segregation, only a minority (12%) of cases have a positive family history, and comparison of case and control allele frequency yielded a point estimate of 8% lifetime risk^7,8^. We observed only 2 deaths past age 90 among V210I individuals, which may not have been due to prion disease (see below), consistent with low penetrance.

The above findings are subject to several biases and caveats. We assumed that individuals for whom we were unable to locate public records of death, such as online obituaries, are still alive. This assumption is supported by the fact that our public record searches were 98.2% sensitive at detecting the deaths of autopsied symptomatic individuals in our dataset. Autopsy cases may represent a biased sample, however, as we also identified individuals who were tested while symptomatic long enough ago that they are very likely deceased, for whom no public records were found. Overall, we estimate that autopsy plus public record searches may be only 92.6% sensitive at detecting deaths. To the extent that we erroneously assumed some individuals to be alive who actually died of prion disease, our estimates may be overly optimistic. On the other hand, we also assumed that deaths identified through public record searches without autopsy were due to prion disease. In fact, particularly for E200K, the range of ages of onset extends into the end of the natural human lifespan. If some deaths were not due to prion disease, then our estimates may be overly pessimistic. The above two limitations act in opposing directions. Sensitivity tests indicate that their potential impact on the analysis may be to shift the median age of onset for E200K a few years in either direction, but regardless, the overall conclusion would still be that E200K has a somewhat later age of onset and lower penetrance than previously believed, while still being highly penetrant.

Diagnosis and ascertainment of older individuals with prion disease appears to be improving over time. For example, U.K. surveillance data indicate that in sporadic prion disease, the measured annual incidence in older individuals, particularly age 80+, was approximately double in 2010-2016 what it was in 1995-2004^13^. Cerebrospinal fluid (CSF) diagnostic tests for prion disease appear to be ordered less often in the elderly, and brain magnetic resonance imaging (MRI) is less sensitive^14^, thus, it is possible that prion disease remains underdiagnosed in older age groups. Our simulations suggest, however, that under-ascertainment of older disease onsets in our snapshot data is not sufficient to explain the younger median onset calculated from those data compared to the longitudinal dataset.

In a previously reported U.K. genetic testing dataset^15^, of 42 individuals who tested positive for a *PRNP* variant, 7/10 who went on to become symptomatic did so within 1 year of testing, a significant excess over expectation. This was interpreted to suggest that some individuals seek predictive genetic testing due to a correct suspicion of initial symptoms of disease. This might introduce bias in the estimation of annual hazards after predictive testing. In contrast, only 3 deaths in our dataset occurred shortly after genetic testing. This difference might be attributable to the composition of genetic variants represented in the two cohorts. Slowly progressive mutations such as P102L, A117V, and 6-OPRI, which are over-represented in the U.K., are more likely to have an extended prodrome as well as more preserved cognition in the earlier stage of the illness, enabling these individuals to seek out predictive genetic testing while possibly experiencing prodromal symptoms. This U.S. cohort consists primarily of rapidly progressive mutations, especially E200K, similar to the world average^7^.

Our study has limitations. No single source of data exhaustively captures all deaths, so our estimate of combined sensitivity is imperfect. We also lacked a perfect gold standard for individuals still alive, thus, we cannot assess the specificity of public record searches. We did not utilize tokenization^16^ or other privacy-preserving record linkage methods to leverage additional sources of vital status information. Only 32 deaths occurred among initially asymptomatic individuals, still a small number from which to estimate hazards. We relied on public obituaries, which are common for ordinary people in the United States, but customs may differ globally, limiting the generalizability of our methods. A strength is that most of our data are from E200K, the most common disease-causing *PRNP* variant worldwide, supporting generalizability of our findings.

The age at symptom onset of genetic prion disease is sufficiently variable that it is not possible to statistically power pivotal pre-approval clinical trials to demonstrate that a drug has delayed onset of disease in this population^2^. Instead, one clinical development path for preventive drugs would require provisional approval on a biomarker endpoint such as cerebrospinal fluid PrP^17,18^, combined with a plan to assess survival long-term in a post-marketing study^19^. Our findings may suggest a need to refine our estimates of annual hazards facing individuals with *PRNP* mutations. An important future direction will be the establishment of a prospective registry study in *PRNP* at-risk individuals. Asking for consent for contact next of kin as well as for tokenization would expand the set of methods available to minimize informative censoring and ensure that no participants are lost to follow-up. Such a study could gather additional prospective data in untreated individuals in advance of prevention trials, while also providing an infrastructure for future post-marketing studies.

## Data Availability

The full de-identified dataset will be made available to qualified investigators with ethical approval and a data use agreement upon request. Binned prospective and retrospective life tables are provided in the Supplementary Materials. These life tables and summary statistics are freely available at github.com/ericminikel/npdpsc_prospective

## Acknowledgments

This study was supported by the National Institutes of Health (R01 NS132022 to EVM), by the Centers for Disease Control and Prevention (NU38CK000486 to BSA) and by donations to the Broad Institute’s Prions@Broad fund. EVM and BSA had full access to all the data in the study and take responsibility for the integrity of the data and the accuracy of the data analysis.

