## Supplemental Material for "Mortality of individuals with antemortem genetic testing for *PRNP* variants in the United States, 1998-2024"

### Supplementary Materials

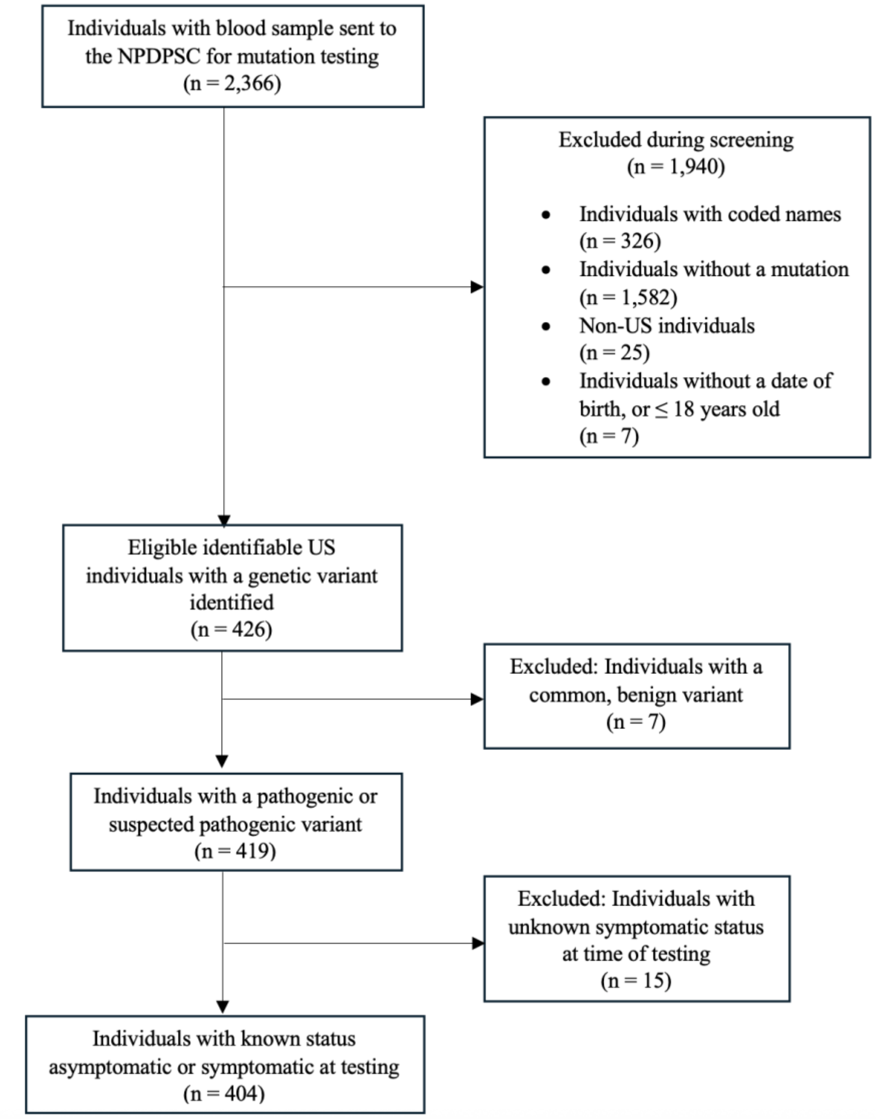


***Figure S1. Participant selection flowchart.*** *This flowchart describes the process of querying and filtering cases from the NPDPSC database for inclusion in this study.*

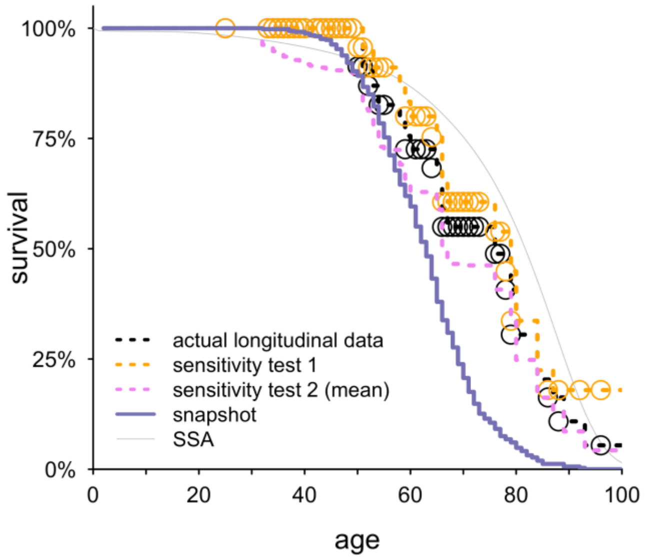


***Figure S2. Sensitivity tests for E200K longitudinal survival curve.*** *Sensitivity test 1: assume that the 4 E200K deaths identified through public records were not due to prion disease, and are treated as censored. This shifts the longitudinal survival curve to the right. Sensitivity test 2: assume that N=3 additional E200K deaths occurred and were not detected by public records nor autopsy. The number N=3 was selected because in symptomatic individuals, the lower bound of the 95% confidence interval on sensitivity of autopsy plus public record searches was estimated to be 87.9% (see Results). 20 E200K individuals asymptomatic at test were known to have died, and if this 20 were to represent a sensitivity of 87.9%, then 2.7 additional deaths went undetected, rounded up to 3. In order to test the most extreme possible assumption, in each of 1,000 bootstrap iterations we randomly selected asymptomatic E200K individuals who were age 31 or older at time of testing, and assumed that they died of prion disease in the same year they underwent testing. The age 31 was selected because it is the earliest ever reported disease onset for E200K out of 571 individuals. Sensitivity test 2 represents the most extreme possible assumption because i) we utilized the lower bound of 95% CI of sensitivity rather than the mean estimate, ii) we assumed that deaths occurred immediately after genetic testing, rather than some number of years later, and iii) we assumed that deaths were uniformly distributed from age 31 onward, when in fact deaths at older ages would be far more likely. The thin violet lines show 1,000 individual bootstrap iterations, and the thick dashed blue line shows the mean survival curve across these iterations. The effect of sensitivity test 2 assumptions is to shift the longitudinal survival curve to the left. Sensitivity tests were then plotted with actual longitudinal data (black), snapshot survival curve, and SSA actuarial curve.*

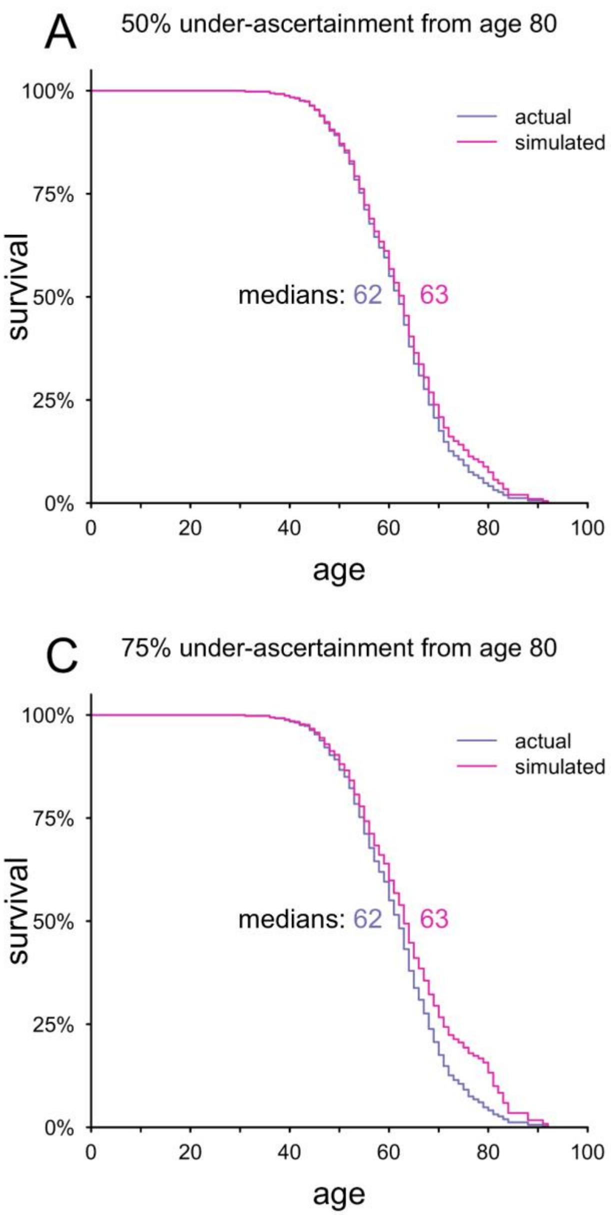


***Figure S3. Simulation of under-ascertainment of older onsets in snapshot data.*** *Using the snapshot life tables for E200K (blue), we simulated what the true survival curve would look like (magenta) in the event that the life tables reflect 50% (A-B) or 75% (C-D) under-ascertainment of deaths over age 80 (A,C) or age 60 (B,D). To do this, we doubled (A-B) or quadrupled (C-D) the number of deaths observed at each integer age at or over the threshold age (80 for A,C and 60 for B,D), and then recomputed the life tables. The number of censored observations in the snapshot life table was not altered.*

| | a) fully naive analysis (neither right- nor left-truncation) | | b) partially naive analysis (right- but not left- truncation) | | c) correct analysis (right- and left-truncation) | |
|---|---|---|---|---|---|---|
| **variant** | **median age of onset** | **penetrance by age 80** | **median age of onset** | **penetrance by age 80** | **median age of onset** | **penetrance by age 80** |
| E200K | 65 | 78% | 83 | 43% | 75 | 69% |
| D178N | 59 | 83% | 61 | 56% | 58 | 76% |

***Table S1. Comparison of naive and correct analyses on longitudinal dataset.*** *For the sake of comparison, we performed naive analyses on our longitudinal dataset, either calculating a) summary statistics for only those individuals who died (no accounting for censoring at all; neither right- nor left-truncation; leftmost columns), or alternatively, b) accounting for censoring of individuals who did not die but not considering the age at entry into the dataset (right- but not left-truncated analysis; center columns). Both naive analyses returned later ages of onset, and lower penetrance, than c) the correct analysis reported throughout this paper (left- and right-truncation). Note that in both (a) and (b) a later age of onset and lower penetrance is observed than in the snapshot data. Note that (a) and (b) are not the correct way to analyze these data, and are presented only for the sake of comparison; only the analysis in (c), which is also the approach used throughout this manuscript, correctly accounts for the ascertainment bias caused by individuals' delayed entry into the dataset at the time of genetic testing. See main Results text for details.*
